# Creating a Structural Resource Prevention Index for Breast Cancer in a Southwestern US City with Historic Neighborhood Segregation

**DOI:** 10.64898/2025.11.26.25341013

**Authors:** Nathan Lothrop, Yoga Korgaonkar, Kimberly L. Parra, Lucia D. Juarez, Violet Wielgus, Francisco E. Mendoza, Celina I. Valencia

**Affiliations:** Department of Community, Environment & Policy, Mel and Enid Zuckerman College of Public Health, University of Arizona, 1295 N. Martin Ave., Tucson, AZ, 85721 USA; School of Geography, Development & Environment, College of Social and Behavioral Sciences, University of Arizona, 1064 E. Lowell Street, Tucson, AZ, 85721 USA; Department of Environmental Health, T.H. Chan School of Public Health, Harvard University, 677 Huntington Avenue, Boston, MA 02115 USA; Heerskink School of Medicine, University of Alabama at Birmingham, Birmingham, Alabama, 35294, USA; College of Education, University of Arizona, 1430 E. Second Street, Tucson, AZ, 85721 USA; Department of Family and Community Medicine, College of Medicine, University of Arizona, 655 N. Alvernon Way, Tucson, AZ, 85711 USA

**Author notes:** Corresponding author: Celina I. Valencia, University of Arizona, 655 N. Alvernon Way, Tucson, AZ, 85711 USA.

## Abstract

**Background:** Historically segregated neighborhoods in the United States continue to experience disproportionate health disparities, including cancer outcomes. The social vulnerability index (SVI) and similar indices are widely used to measure community disadvantage, but they do not directly capture contextual infrastructure relevant to cancer prevention. In Tucson, Arizona, where racially restrictive housing covenants, conditions, and restrictions (CCRs) remain as a legacy of segregation, we developed an alternative approach to assess neighborhood-level access to breast cancer prevention resources.

**Methods:** We developed the Structural Prevention Resources Index (SPRI), calculated as arithmetic mean of scores for access to free screening clinics, grocery stores with fresh produce, and parks. Each component was equally weighted to produce a composite score ranging from 1 (better access) to 6 (poorer access). We compared the SPRI to the Social Vulnerability Index (SVI).

**Results:** Drive-time analyses highlighted poor access to clinics providing free breast cancer screening for the eastern study area. Park access and quality were more evenly distributed with 56% of women ages 40-64 having access to high-quality parks. SPRI showed that minoritized women ages 40-64 more often resided in high or very high resource areas (67%) vs. non-minoritized women ages 40-64 (56%). SPRI captured geographic variation distinctly different than the SVI, notably a large central eastern portion of the study area with few prevention resources and low SVI scores.

**Conclusion:** Disease-specific indices such as SPRI can uncover structural barriers to chronic disease prevention, like breast cancer, often overlooked by general measures of geospatial vulnerability, offering more precise tools for guiding equitable public health planning and strategies.

**Highlights:**

- Develops the Structural Prevention Resource Index (SPRI) for breast cancer.
- Compares disease-specific index (SPRI) with the Social Vulnerability Index (SVI).
- Finds SPRI captures geographic disparities not reflected in SVI in Tucson, Arizona.
- Demonstrates value of disease-specific indices for public health planning.

## 1. Introduction

In the United States (US), breast cancer accounts for 30% of all new female cancers per year, killing over 40,000 women annually [1]. Meanwhile, disparities in breast cancer outcomes and negative quality of life for survivors not only persist but continue to widen [2,3]. Black or African American, American Indian or Alaska Native, Asian, Native Hawaiian and Other Pacific Islander and Hispanic/Latina women, henceforth referred to as minoritized women, have lower overall incidence rates of breast cancer than non-Hispanic White (NHW) women. However, when minoritized women do receive a diagnosis, they are more likely diagnosed at a later disease stage and more likely to die from the disease [1,2,4]: an epidemiological phenomenon termed the breast cancer mortality disparity. Over the last 10 years, breast cancer not only remains the leading cancer burden among women in the U.S. but is accompanied by increasing incidence rate and steep rise in diagnosis before the age of 50 [1].

To improve cancer outcomes across the continuum, evidence-based recommendations suggest a diet high in fruits and vegetables, engaging in consistent leisure time physical activity, and maintaining a healthy weight [5]. Early detection of breast cancer is associated with improved outcomes. The US Preventive Services Task Force recommends initiating routine mammography screening at age 40 and continuing every two years until age 74 [6]. These recommendations are primarily discussed in the context of breast cancer prevention but also increase positive survivorship outcomes [7]. To better understand trends of breast cancer disparities, it is critical to understand how access for minoritized women to health resources and opportunities for prevention is shaped by social positionality.

Neighborhood environments are widely recognized as powerful determinants of health [8], with extensive literature linking socioeconomic, physical, and social characteristics of place to health outcomes, specifically area-level morbidity and mortality across the life course. However, the tools that have been commonly employed to capture neighborhood constructs like neighborhood deprivation [9,10] or composite social vulnerability [11–14] were not designed with health prediction as their primary aim. Rather, these measurement tools were developed to understand resource allocation, policy planning, or broad population surveillance. As a result, their ability to capture disease-specific risk pathways is dampened. To address this gap, we introduce the Structural Prevention Resource Index (SPRI), a disease-specific index of contextual access to prevention resources. We tailored neighborhood measures specific to breast cancer directly; our approach represents an important methodological advancement that sharpens the link between place and disease risk, offering a more precise tool for epidemiological and translational work.

Redlining as a driver of breast cancer disparities has been previously considered in the literature [8]. Current residents of neighborhoods that were “redlined”, a federal practice originating in the 1930s of categorizing areas with higher shares of racial or ethnic minority residents as financially riskier for making home loans, also experience increased breast cancer rates [11–13]. While this practice was common in established cities at the time, smaller ones had limited or no redlining [14]. Historical documents attest to most cities participating in some form of residential segregation maintained by racially restrictive housing covenants, conditions, and restrictions (CCRs) [15]. While these CCRs were rendered unenforceable in 1948, they were not illegal until the Fair Housing Act (FHA) of 1968 [16]. Even after the enactment of the FHA, historically segregated neighborhoods continued to experience disinvestment, thereby influencing contemporary access to health in areas with high percentages of minoritized residents [17,18].

Our study focuses on Tucson, Arizona, a unique city to consider the complexities and contemporary remnants of historical residential segregation, that did not participate in formal redlining. Arizona segregation laws were among some of the most stringent for Blacks and Latinos prior to the FHA [19]. In Tucson, political, economic, and ideological forces, along with enacted policies, resulted in segregated ethnic enclaves known as barrios [17]. The enduring legacy of Tucson’s residential segregation can also be tracked through legal struggles surrounding public school desegregation. Beginning in 1973, the city’s largest school district, Tucson Unified School District (TUSD), was put under court supervision for failing to achieve school desegregation [20], which was only lifted in 2022 [21]. The totality of these social processes demonstrates that artifacts of historical segregation may inform the current built environment, potentially increasing health disparities, notably breast cancer. We are unaware of any study that has previously examined the built environment for relationships between historic residential segregation and present-day disease prevention resource access at the neighborhood level.

In this paper, we will examine racial CCRs on neighborhood-level breast cancer prevention infrastructure. We will develop the SPRI, based on potential access to prevention resources. The SPRI measures access to: 1) free routine cancer screening, specifically mammography under the National Breast and Cervical Cancer Early Detection Program (NBCCEDP); 2) healthy food options; and 3) parks or green spaces that provide potential access to physical activity [5]. The SPRI provides a comprehensive representation of the enduring historical legacy of residential segregation to understand contemporary patterns of breast cancer prevention disparities experienced by minoritized women via potential access [22]. To our knowledge, this is the first study to center disease-specific prevention health resource access within the context of contemporary built environments shaped by historic residential segregation outside of redlining.

## 2. Methods

To examine the relationship between areas with historic racial CCRs and access to breast cancer risk reduction infrastructure (e.g., fresh food, parks), we developed a multistep model that uses the most recent publicly available spatial data. The key steps include:

1. Calculating SPRI scores for individual domains of breast cancer risk reduction resources.
2. Calculating SPRI score for census blocks; and
3. Analyzing the relationship between residentially segregated areas and SPRI scores.

### 2.1 Study Setting

This study focuses on the Tucson metropolitan region in Pima County, Arizona, USA (Figure 1), including the cities of Tucson and South Tucson, as well as portions of the towns of Marana to the northeast, Oro Valley to the north, Sahuarita to the south, and unincorporated areas of eastern Pima County. The study area encompasses 12,474 census blocks that fall within these jurisdictions, representing a total population of 922,696, of whom 466,774 (50.59%) are identified as people of color and 237,927 (25.79%) as minoritized women.

**Fig. 1.**
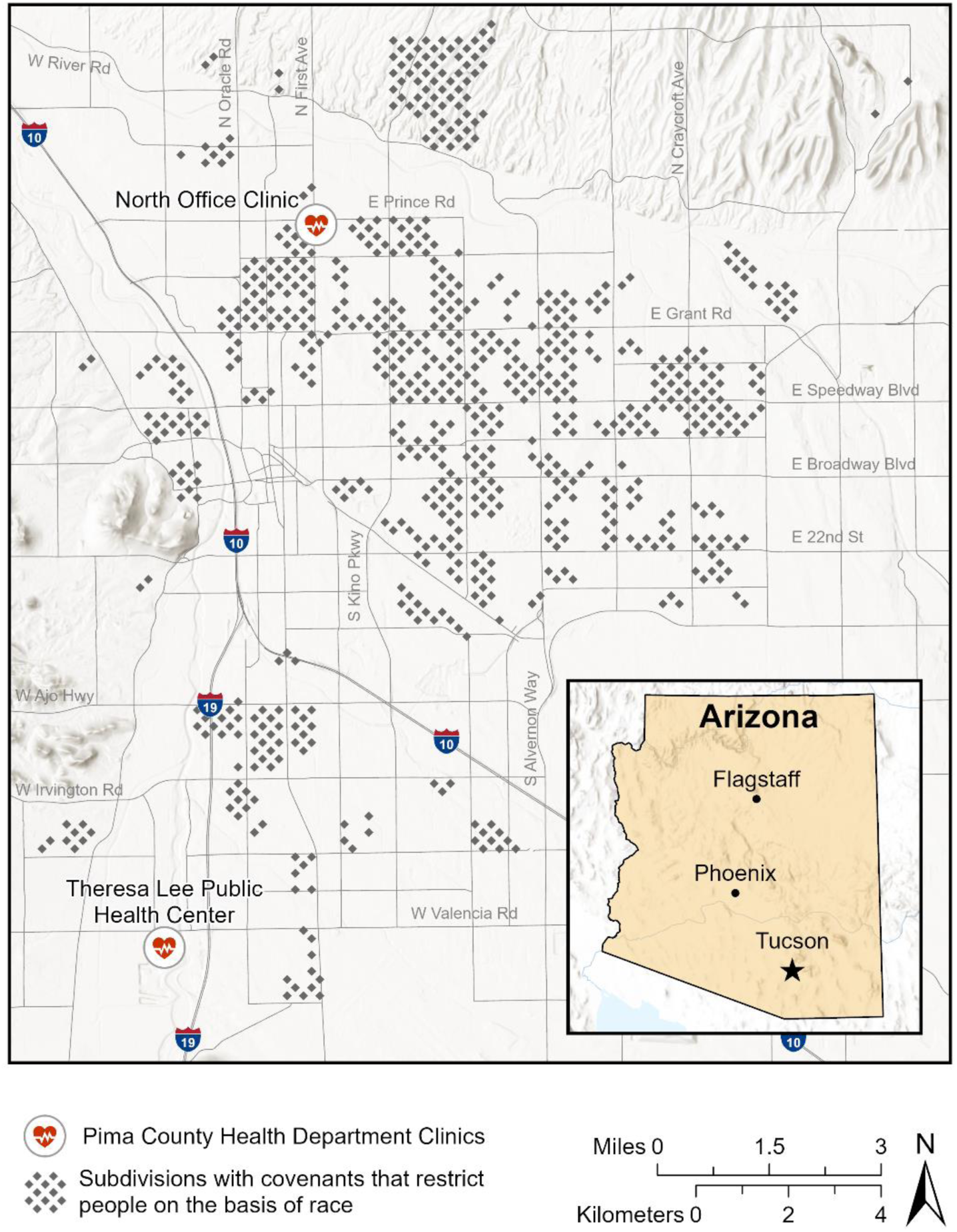
Locations of clinics that offer free mammography and other preventative services and subdivisions with racial CCRs in the study area.

### 2.2 Data

#### 2.2.1 Racial CCRs and Demographics

Data on study area residential subdivisions containing racist covenants were obtained from the Mapping Racist Covenants Project [22]. Briefly, the Mapping Racist Covenants project compiled this data using historic public records from the Pima County Recorder’s Office and verified the data for the presence of racist language. This dataset includes 1,338 subdivisions recorded between 1912 and 1968 that contained racist language restricting certain racial groups from owning or renting properties in specific areas of Tucson (Figure 1). As many as 209 subdivisions excluded people based on their race/ethnicity. Specifically, 169 subdivisions enacted “Caucasian or White Only” clauses, 145 subdivisions explicitly excluded Black/African American individuals, and 142 subdivisions explicitly excluded Asian individuals. We obtained demographic data for 12,474 census blocks from the 2020 US Decennial Census [23].

#### 2.2.2 Breast Cancer Prevention Infrastructure

The Pima County Health Department offers free breast cancer screenings through the Well Woman Health Check program, with services including but not limited to breast exam, mammography, biopsy, referral for treatment, case management, and translation [24]. Two health clinics operate year-round to offer free screenings: Teresa Lee Public Health Center and North Office Clinic. Additionally, five other locations managed by El Rio Health are also part of this program but were omitted from this analysis as these sites only offer free screenings during October, as it is Breast Cancer Awareness Month. Consequently, only the two clinics operated by Pima County Health were considered for this study (Figure 1).

For data on locations of fresh food vendors, we included grocery stores in the study area that stock fresh produce and accept Women, Infants, & Children (WIC) and Supplemental Nutrition Assistance Program (SNAP) benefits [25], as well as farmers markets that were part of the Arizona Farmers Market Nutrition Program [26]. Our dataset included 70 grocery stores and 4 farmers markets, further referred to as grocery stores. As a physical activity proxy, we used two datasets to represent public parks and green spaces. The first is the Pima County Parks and Recreational Areas dataset of 311 parks [27], augmented by quality scores based on amenities such as play areas, ramadas, water fountains, park size and vegetation types [28]. The second dataset was of park entrances to more accurately represent accessibility [28].

### 2.3 Analysis

#### 2.3.1 Calculate Access Index Scores for Breast Cancer Risk Reduction Infrastructure

Service area analysis, a component of network analysis, identifies the geographic area that can be reached within a specified distance or travel time along a road network, accounting for road connectivity, travel costs and constraints, and real-world accessibility. Using the service area solver in drive time mode in the Network Analyst extension of ArcGIS (ESRI, Redlands, CA), we created service area polygons for health clinics, fresh food vendors, and green space entrances at ≤2, 2-5, 5-10, 10-15, 15-30 and >30-minute drive times. The driving times from the service area analysis were used to create an accessibility index for each census block, representing access to breast cancer risk reduction infrastructure. Index values ranged from 1 for a 2-minute drive to 6 for a >30-minute drive. Walking analysis was not conducted as the Tucson metro area is car-centric and walking during heat season may be dangerous or even fatal.

#### 2.3.2 Augmenting Park Service Area with Amenities Index Scores

In addition to driving time, each park was assigned an amenity score ranging from 0 to 6, calculated by summing the scores for any play areas, ramadas, water fountains, or restrooms with park size and vegetation type. A score of 0 represented no amenities, sparse vegetation and smaller park size as compared to a score of 6 that represented the presence of all amenities, dense vegetation and a large park size. The Spatial Join tool in ArcGIS (ESRI, Redlands, CA) was used to assign the parks’ drive times to each census block using a one-to-many relationship because each census block had access to multiple parks with different drive times. For each census block, a park index was calculated by dividing the park amenity score by drive time [29], with results ranging from 0 to 3. A value of 0 represents a high drive time and low park amenity score, whereas a value of 3 represents a shorter drive time and a high park amenity score. This index was used to filter parks with shorter drive times and high amenities score for each census block. After filtering the parks, the park accessibility index was scaled to a range of 1 to 6 for consistency with other accessibility indices, where a value of 1 indicates high accessibility with shorter drive times to parks that have better amenities, denser vegetation, and larger park sizes. Conversely, a value of 6 represents low accessibility, indicating longer drive times to parks with fewer amenities, sparse vegetation, and smaller park sizes.

#### 2.3.3 Calculating Structural Prevention Resource Index Scores

The SPRI score as an indicator for disease prevention resources was calculated at the census block level from the arithmetic mean of the access scores for screening clinics, grocery stores, and parks, resulting in a score ranging from 1 to 6. A score in the range of 1-2 indicates better access to health clinics, grocery stores, and parks (lower breast cancer risk), and a score in the range of 5-6 indicates poorer access to resources (and higher risk).

#### 2.3.4 Comparison to Social Vulnerability Index

The Social Vulnerability Index (SVI) was created by the Geospatial Research, Analysis, and Services Program (GRASP) to identify communities experiencing social vulnerability before, during or after natural disasters [30]. The current SVI combines 16 U.S. census variables, divided into four themes of 1) Socioeconomic Status, 2) Household Characteristics, 3) Racial & Ethnic Minority Status, and 4) Housing Type & Transportation, into a single index of social vulnerability at the census tract scale. SVI data was downloaded for the year 2022 [31] and was used to compare areas of vulnerability with the SPRI. The SVI, while not constructed for the purpose of understanding aggregate neighborhood-level health outcomes has been increasingly used for epidemiological studies. In cancer research, the SVI has been applied to evaluate the influence of neighborhood-level social and structural determinants on breast cancer screening [32], treatment patterns [33], and survival outcomes [34]. Additionally, SVI for cancer incidence rates have been found to have contradictory findings [35].

Human subjects research approval was not required for this ecological study as we did not involve any human subjects or data related to identifiable individuals. All spatial datasets were projected to the NAD 1983 State Plane Arizona Central FIPS 0202 (Intl Feet) projected coordinate system (EPSG 2223) to ensure consistency across datasets during analysis in ArcGIS (ESRI, Redlands, CA).

## 3. Results

### 3.1 Service Area Analysis

The drive time service areas map for health clinics highlights a significant lack of accessibility to health clinics offering free breast cancer screening services in the eastern part of the study area, with drive times >15 minutes in these areas (Figure 2a). Drive time service areas for grocery stores reveal the presence of food deserts in the western part of the study area, as well as a smaller area in the southeast where drive times exceed 5 minutes to the nearest grocery store (Figure 2b). However, much of the study area is within a 15-minute drive to the nearest grocery store that offers fresh produce and accepts WIC and SNAP benefits. Parks are well distributed throughout the study area, seldom more than a 5-minute commute (Figure 2c).

**Fig. 2.**
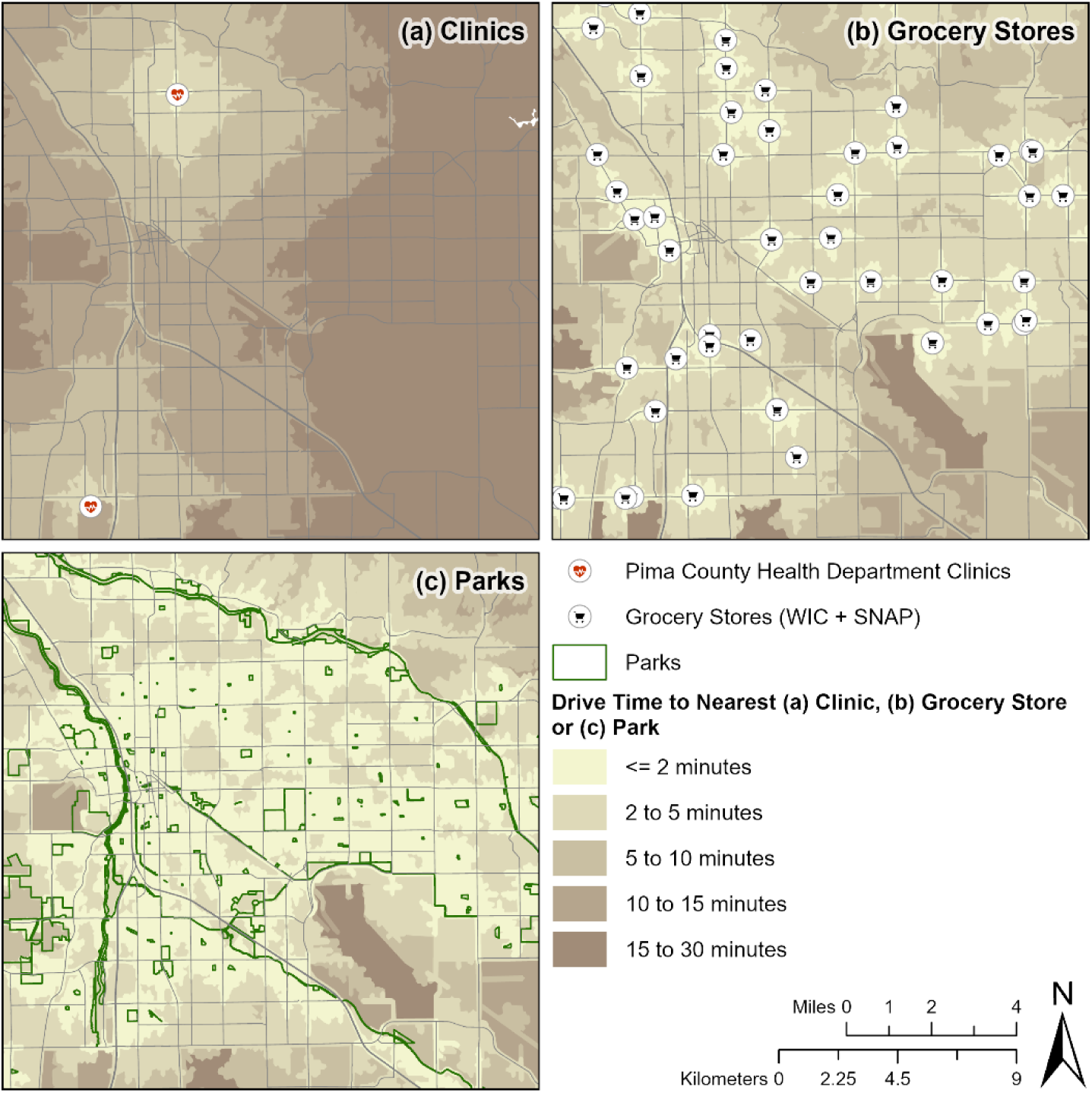
Drive times to the nearest (a) clinic, (b) grocery store, and (c) park for each census block.

### 3.2 Park Access and Quality

For park access and quality, we found a more nuanced picture of this component of breast cancer risk reduction. Approximately 56% of women ages 40-64 years have access to high or very high-quality parks, and this percentage is nearly identical to other demographic groups (Table 1).

**Table 1.**
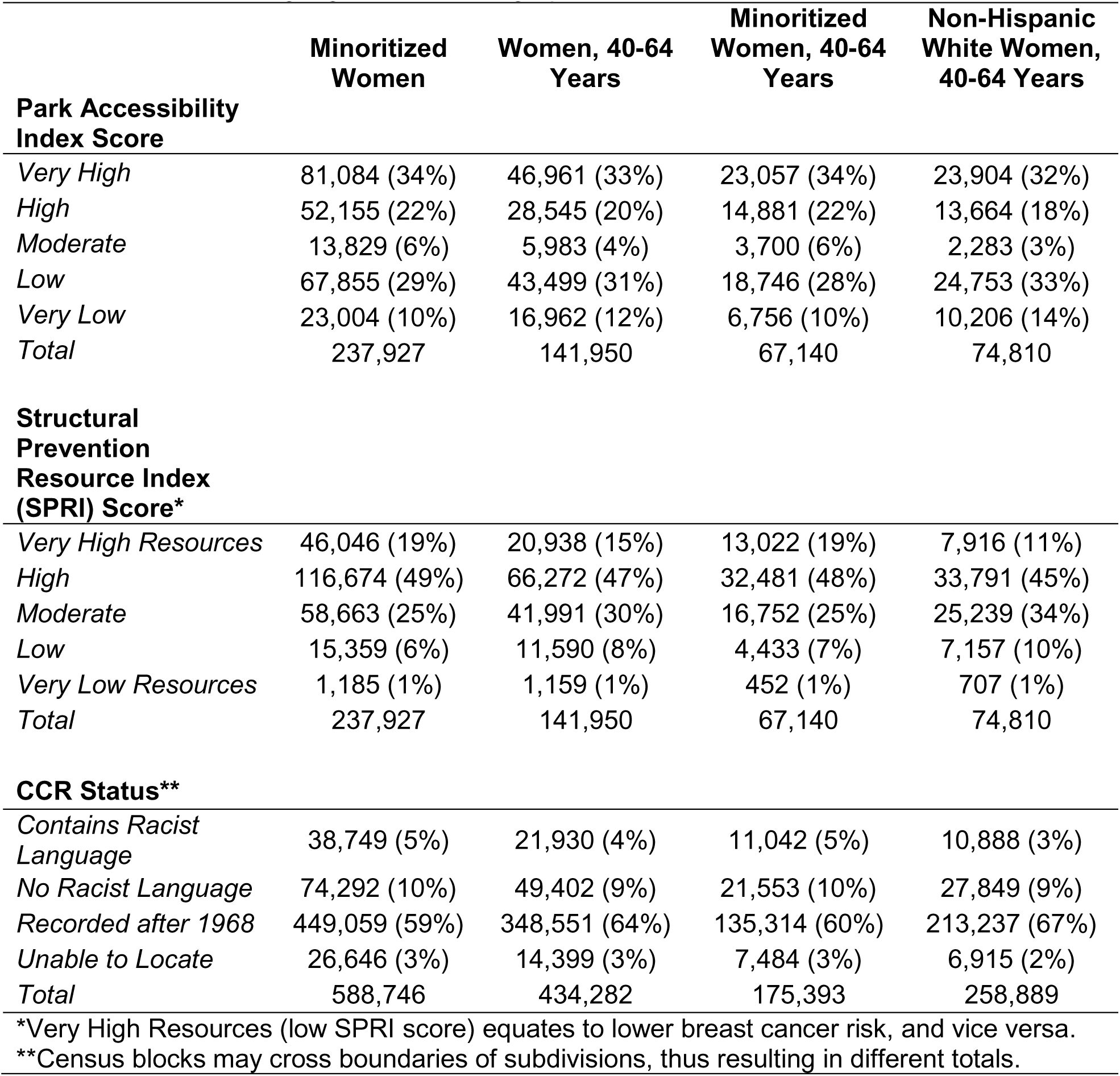
Park access and Structural Prevention Resource Index (SPRI) scores and racially restrictive covenant language across demographics.

|  | Minoritized Women | Women, 40-64 Years | Minoritized Women, 40-64 Years | Non-Hispanic White Women, 40-64 Years |
| --- | --- | --- | --- | --- |
| **Park Accessibility Index Score** |  |  |  |  |
| *Very High* | 81,084 (34%) | 46,961 (33%) | 23,057 (34%) | 23,904 (32%) |
| *High* | 52,155 (22%) | 28,545 (20%) | 14,881 (22%) | 13,664 (18%) |
| *Moderate* | 13,829 (6%) | 5,983 (4%) | 3,700 (6%) | 2,283 (3%) |
| *Low* | 67,855 (29%) | 43,499 (31%) | 18,746 (28%) | 24,753 (33%) |
| *Very Low* | 23,004 (10%) | 16,962 (12%) | 6,756 (10%) | 10,206 (14%) |
| *Total* | 237,927 | 141,950 | 67,140 | 74,810 |
| **Structural Prevention Resource Index (SPRI) Score\*** |  |  |  |  |
| *Very High Resources* | 46,046 (19%) | 20,938 (15%) | 13,022 (19%) | 7,916 (11%) |
| *High* | 116,674 (49%) | 66,272 (47%) | 32,481 (48%) | 33,791 (45%) |
| *Moderate* | 58,663 (25%) | 41,991 (30%) | 16,752 (25%) | 25,239 (34%) |
| *Low* | 15,359 (6%) | 11,590 (8%) | 4,433 (7%) | 7,157 (10%) |
| *Very Low Resources* | 1,185 (1%) | 1,159 (1%) | 452 (1%) | 707 (1%) |
| *Total* | 237,927 | 141,950 | 67,140 | 74,810 |
| **CCR Status\*\*** |  |  |  |  |
| *Contains Racist Language* | 38,749 (5%) | 21,930 (4%) | 11,042 (5%) | 10,888 (3%) |
| *No Racist Language* | 74,292 (10%) | 49,402 (9%) | 21,553 (10%) | 27,849 (9%) |
| *Recorded after 1968* | 449,059 (59%) | 348,551 (64%) | 135,314 (60%) | 213,237 (67%) |
| *Unable to Locate* | 26,646 (3%) | 14,399 (3%) | 7,484 (3%) | 6,915 (2%) |
| *Total* | 588,746 | 434,282 | 175,393 | 258,889 |
| *Very High Resources (low SPRI score) equates to lower breast cancer risk, and vice versa. |  |  |  |  |
| **Census blocks may cross boundaries of subdivisions, thus resulting in different totals. |  |  |  |  |

However, there were distinct spatial patterns of park access, where large areas in the central eastern and southern study area have markedly lower quality and access scores as shown by the dark green color (Figure 3). When comparing the drive time service areas to the nearest parks (Figure 2c), despite being close to parks with drive times less than 5 minutes, many areas of Tucson face limited access to higher quality parks.

**Fig. 3.**
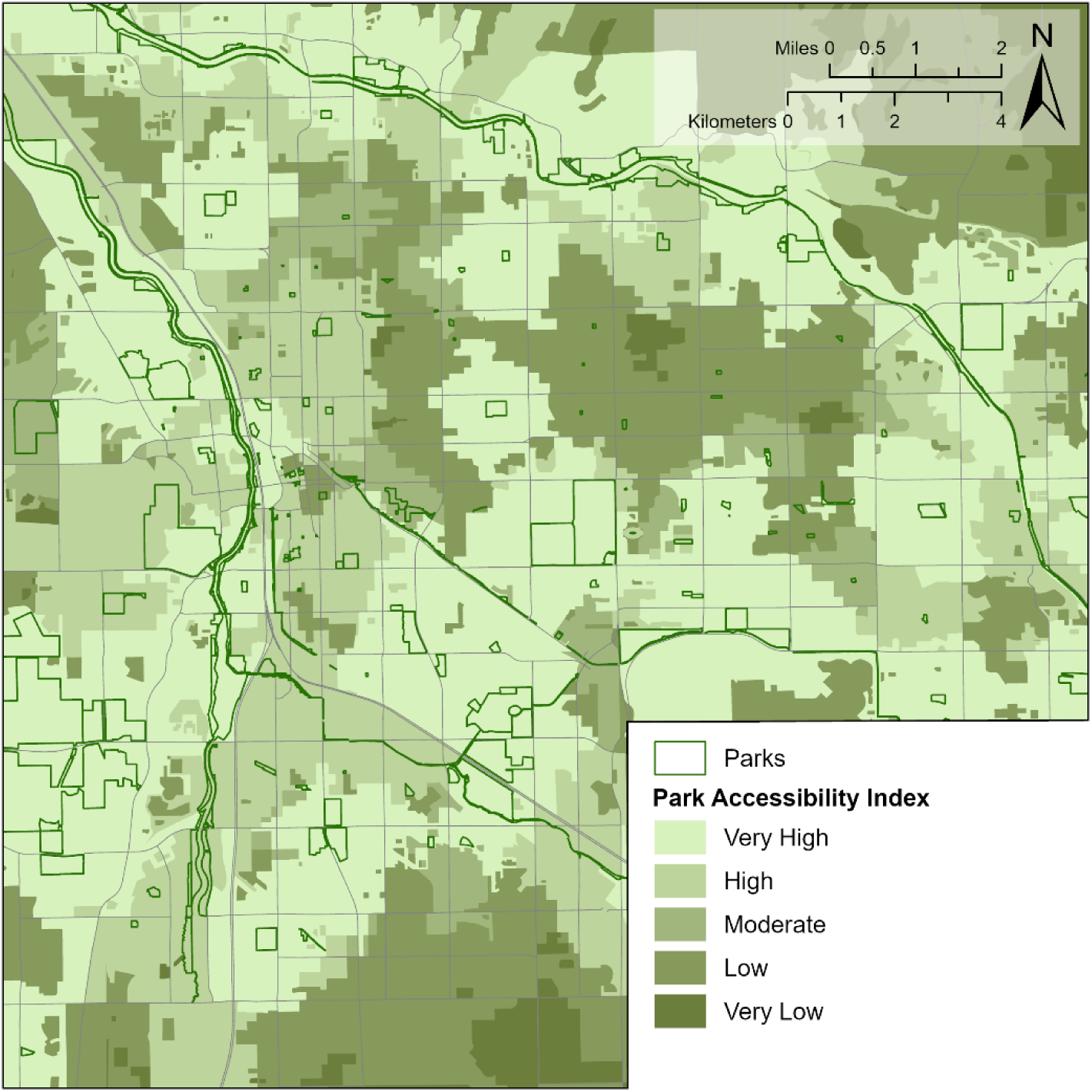
Park quality and access index mapped for the study area. Darker shades of green indicate longer drive times, fewer amenities, less vegetation, smaller park sizes; and vice versa for the lighter shades of green.

### 3.3 Structural Prevention Resource Index (SPRI)

The SPRI combines access or drive time scores for clinics and grocery stores with park access and quality scores to geographically based risk. We are conceptualizing risk as the reduction of access for health resources and opportunities to prevent breast cancer. Overall, minoritized women ages 40-64 years lived in higher resource (i.e., lower risk) areas compared to NHW women of the same age (Table 1). Among minoritized women ages 40 to 64, 67% live in high or very high resource areas, more than NHW women of the same age (56%). Similarly, there were 9% more NHW ages 40-64 in moderate resource areas versus minoritized women in this age bracket.

Similar to the spatial patterns seen in Figure 3, there are large areas with moderate resources in the central east and southern parts of the study area, punctuated by pockets of low resource scores (Figure 4). Low and very low resource areas ring the study area. There are no visual geographic patterns of subdivisions with racially restrictive language and breast cancer risk (Figure 4).

**Fig. 4.**
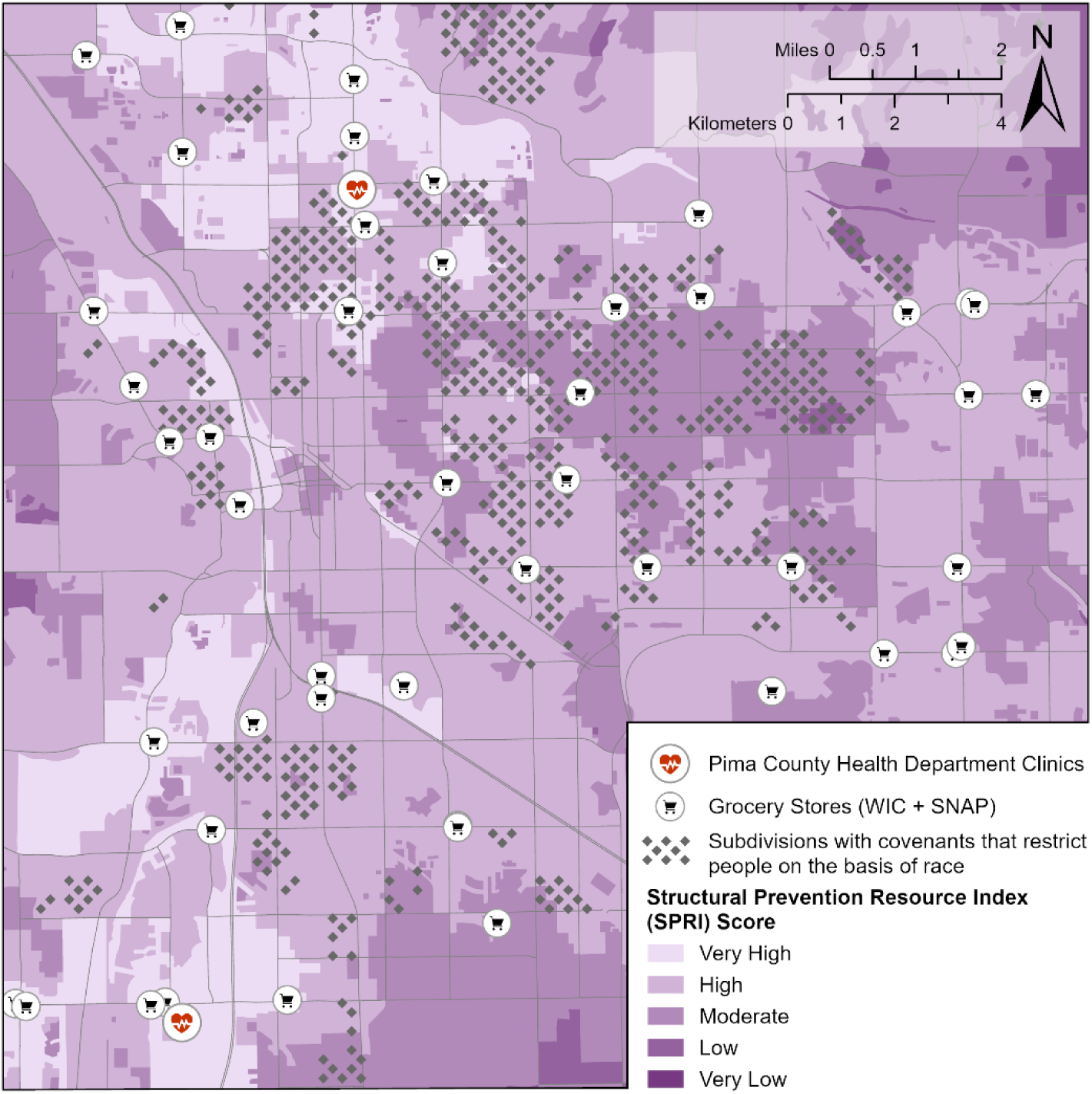
Structural Prevention Resource Index (SPRI) scores along with subdivisions with racially restrictive language, clinics, and grocery stores.

In comparing racially restrictive CCRs among demographic groups, we found minoritized women in middle-age tended to live in areas with CCRs recorded after 1968 (60%), with only 5% living in subdivisions with racially restrictive language (Table 1), with no meaningful distinctions among NHW women.

There were small differences in SPRI scores across subdivisions by racially restrictive language category, in which women living in areas with very high or high SPRI scores (i.e., lower breast cancer risk) were less likely to have the language recorded after 1968 (Table 2), indicating more resources tended to be in older subdivisions. However, this should be interpreted with caution given the small numbers of women in areas with racist language and low or very low resources (n=51).

**Table 2.** Total women ages 40-64 by Structural Prevention Resource Index (SPRI) score and racially restrictive covenant language categories.

| Structural Prevention Resource Index (SPRI) Score* | Racially Restrictive Covenant Language Status |  |  |  |  |
| --- | --- | --- | --- | --- | --- |
|  | *Contains Racist Language* | *No Racist Language* | *Recorded after 1968* | *Unable to Locate* | *Total\*\** |
| *Very High Resources* | 6,771 (10%) | 11,182 (17%) | 44,651 (67%) | 4,357 (7%) | 66,961 |
| *High* | 11,044 (5%) | 25,318 (12%) | 162,444 (79%) | 7,368 (4%) | 206,174 |
| *Moderate* | 4,064 (3%) | 11,310 (9%) | 114,328 (87%) | 2,358 (2%) | 132,060 |
| *Low* | 51 (<1%) | 1,017 (4%) | 26,121 (95%) | 213 (<1%) | 27,402 |
| *Very Low Resources* | 0 | 575 (34%) | 1,007 (60%) | 103 (6%) | 1,685 |

\*Very High Resources (low SPRI score) equates to lower breast cancer risk, and vice versa.
\*\*Census blocks may cross boundaries of subdivisions, thus resulting in different totals.

### 3.4. SPRI Comparison to SVI

The 2022 SVI indicates higher social vulnerability in the north-western and south-western regions of the study area (Figure 5). This is in sharp contrast to the SPRI scoring, as these same areas have high or very high resources scores (Figure 4). High SVI and low SPRI scores do overlap in the eastern central portion of the study area.

**Fig. 5.**
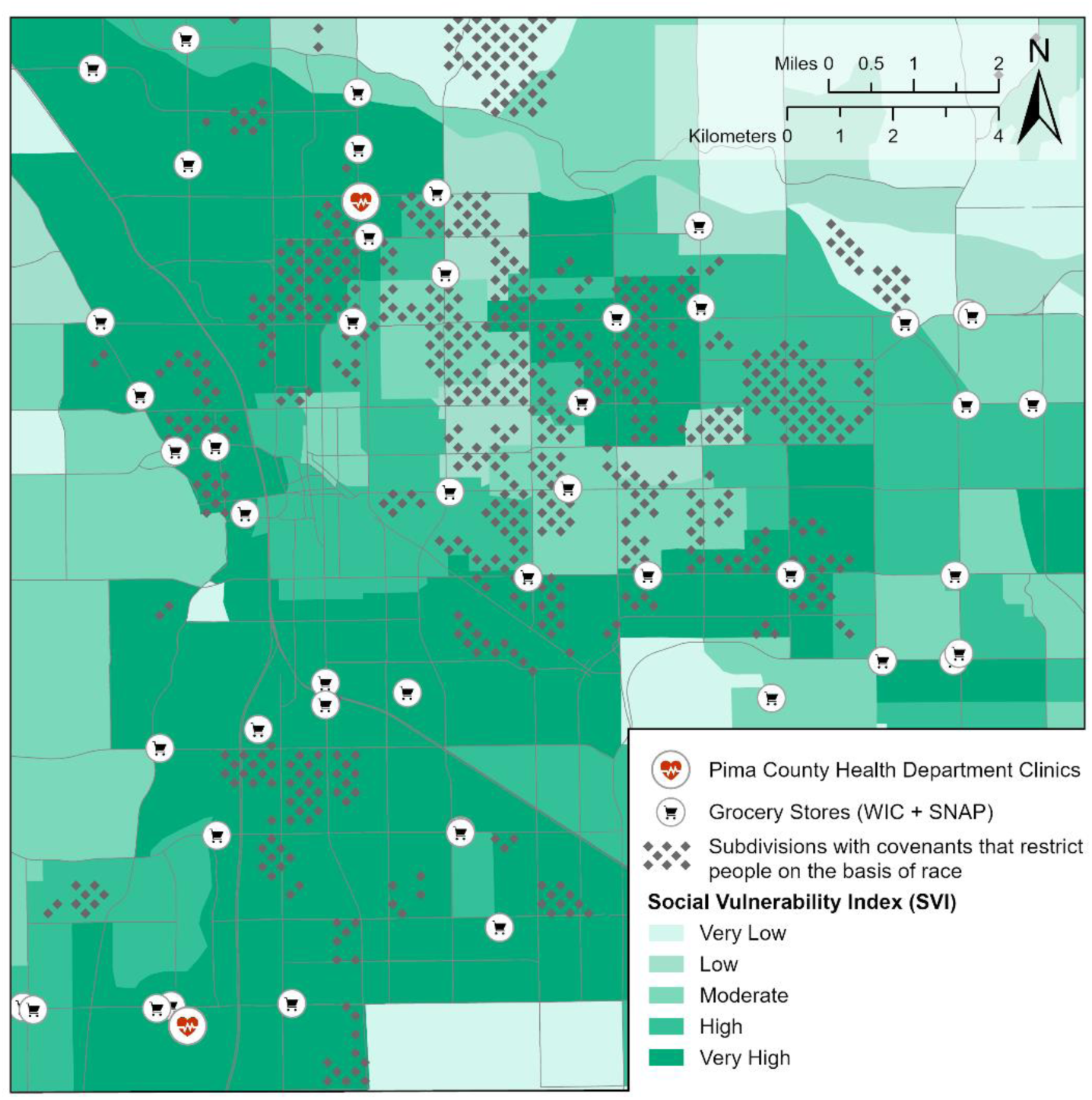
Social Vulnerability Index (SVI) along with subdivisions with racially restrictive language, clinics, and grocery stores.

## 4. Discussion

To our knowledge, this is the first study to build a disease-specific spatial framework for prevention and access findings in the context of racial CCRs in a city that did not participate in formal redlining practices. We found some differences across demographic groups and by historic racial CCRs, as well as the geographic distribution of breast cancer risk across the study area. NHW women were slightly less likely to live in an area with racially restrictive covenants compared to minoritized women, though the vast majority of all women lived in areas without any racialized land use language. Minoritized women ages 40-64 tended to live in lower breast cancer risk neighborhoods compared to their NHW counterparts.

Our findings indicate that while the SPRI shares some overlap with SVI, there are vast portions of the study area with contradictory findings (Figures 3 and 4). The limited agreement suggests that both indices capture broad structural determinants of health, yet the divergence underscores that a disease-specific index may identify pathways more directly relevant to cancer risk outcomes. This distinction is important as the SVI was designed as a general measure of social vulnerability [36], whereas our tailored approach integrates features more germane to cancer epidemiology. While there are similar indices at census block-group [10] and tract level [9], they use the same number or fewer of very similar census variables, yet are also disease agnostic and do not differentiate by sex. The SPRI may be more representative of the geographic patterns of disease, with increased sensitivity to inequities in cancer prevention, diagnosis, and care.

Out of the 16 U.S. census variables used in the creation of SVI, only certain variables are relevant for assessing breast cancer risk. The variables in the socioeconomic theme are not directly related to breast cancer risk. The household characteristics theme includes two age related variables, “Aged 16 & older” and “Aged 17 & younger,” but they do not account for the specific age group most at risk among women. The racial & ethnic minority status theme includes variables that could potentially inform such risk by identifying minoritized populations, but without distinction by sex. Finally, the housing type & transportation theme does include a “no vehicle” variable that could potentially be used as a proxy for accessibility to healthcare, grocery stores or recreation. Overall, however, the SVI includes multiple variables that have little or no influence on breast cancer prevention or risk. By contrast, the SPRI is designed to incorporate factors that directly tie to breast cancer prevention, making it a more precise and appropriate tool for identifying areas with limited resources to reduce breast cancer risk.

Based on previous literature, we anticipated that minoritized women would live in areas with higher breast cancer risk [37,38] and in areas with historic racial CCRs that signal residential segregation [39]. Contrary to national patterns, SPRI indicated that minoritized women ages 40-64 disproportionately lived in better accessed areas compared to NHW women, reflecting Tucson’s distinctive urban geography. These differences reflect the utility of disease specific indices, which may reveal locally contingent structural dynamics that may not be evident in previous studies that have generally relied on neighbor deprivation or social vulnerability indices [40]. Our method does not use disease-agnostic data like social vulnerability indices, as these indices are related to breast cancer outcomes [41] and screening rates [42], as well as innumerable other outcomes [40]. While such analyses may illuminate relationships between social factors and disease outcomes, they can frame neighborhoods in irreparable deficit without actionable solutions. Importantly, these analyses also are devoid of neighborhood perceptions that may dampen some features of the built environment that have been approached in the literature to be deprivation or vulnerability [43]. Future directions include applying SPRI to urban areas that used historical redlining practices to assess if the residential segregation strategy used in Tucson are distinct and have resulted in unique health resource distribution patterns across the area.

Another explanation is that public prevention infrastructure is suited to generally serve all demographic groups in the city. In the most recent City of Tucson Parks and Recreation Master Plan from 2016, neighborhood and community parks were the highest priority for funding, which would direct funding to more equitable distribution of parks across the study area [44] (Figure 2c), though these vary in quality (Figure 3). Clinic locations are highly concentrated in the City of Tucson (Figure 2a). Should there be the opportunity to expand the program, we suggest researching current and potential capacity, whether at brick and mortar or mobile clinics. Capacity and utilization patterns can vary widely [45], and the Well Woman Health Check Program, which is funded through the Arizona Health Care Cost Containment System, may be severely restricted by H.R.1 (the “One Big Beautiful Bill Act”) [46].

Interestingly, minoritized women were only slightly more likely to live in areas with historic racial CCRs, far less that what might be expected based on redlining, which did not occur in the Tucson area [39]. We are unaware of any other studies that have investigated this form of structural racism in relationship to a health risk, much less in the context of a city along the US-Mexico border with a complex history of its own, and which had a population of 32,506 people in 1930 [47], around when redlining began in more populous cities. While we do not dispute structural racism, future studies could investigate locations of Latino ethnic enclaves (i.e., barrios) [48] and contemporary gentrification of these neighborhoods that may influence neighborhood demographics to understand their influence on breast cancer risk.

Breast cancer risk, as a function of less access to prevention resources and opportunities, was higher in central eastern and south-central study area neighborhoods due to limited prevention infrastructure (Figures 2a-c and 3). Our study does not account for tract-level data, notably personal vehicle access, which could modify how easily preventative infrastructure could be reached. However, 97% of study area residents have access to at least one vehicle for daily use [49], which may make this point moot. The increased risk on the outskirts of the Tucson metropolitan area may result from sparse population and resulting fewer grocery stores and parks to serve this population, which echoes prevention resource access issues in increasingly less populated areas [50]. However, we also identified a large area of high breast cancer risk in the eastern central portion of the metro area within the boundary of the City of Tucson, differentiated from the rest of the city by total lack of grocery stores and poor park access and quality (Figures 2b and 3). Future directions will be to analyze SPRI scores in relation to cancer outcomes to test predictive capacity and validity of the tool.

We relied on publicly available data at the neighborhood scale to produce a fine-spatial resolution mapping of prevention infrastructure that, except for grocery stores, relies on free preventative infrastructure that directly relates to the disease of interest. While such prevention data at neighborhood scale may limit adaptation beyond our area, other studies typically utilize individual study data, which are not available for our area and likely not for many others. Another limitation of our approach is the mismatch spatial boundaries between census block and neighborhood boundaries. Although area-weighted averaging could be used under the assumption that census variables are uniformly distributed across each census block or neighborhood, we did not employ this method and are unaware of alternative approaches that would notably improve accuracy.

Other studies tend to use census tract or zip code level spatial resolution with preference to screening access and/or capacity [45,51–54]. While these approaches are important, they may not be practically useful for local public health officials at city or county levels due to coarse spatial resolution or not practical to implement due to data availability, technical capacity, or timing constraints. In addition, by focusing only on clinic service area capacity from a public health perspective, this may perpetuate isolating public health offices from transportation, parks and recreation, and planning offices, which are crucial to engage to develop a more geographically complete cancer prevention framework, as illustrated by our approach. This is critically important, as effective public health policy interventions at the US federal level seem increasingly unlikely, placing a greater onus of prevention on state, county, and local governments.

## Author Statement

### Author contributions: CRediT Author contributions: CRediT

**Nathan Lothrop**: Writing – review and editing, Writing – original draft, Methodology, Investigation.

**Yoga Korgaonkar**: Writing – review and editing, Visualization, Methodology, Investigation, Formal analysis, Data curation.

**Kimberly L. Parra**: Writing – review and editing, Writing – original draft, Conceptualization, Methodology.

**Lucia Juarez**: Writing – review and editing, Conceptualization, Methodology.

**Violet Wielgus**: Writing – review and editing, Visualization, Methodology, Investigation, Data curation.

**Francisco E. Mendoza**: Writing – review and editing, Methodology, Investigation, Conceptualization.

**Celina Valencia**: Writing – review and editing, Writing – original draft, Supervision, Methodology, Investigation, Conceptualization, Funding acquisition.

## Data Availability

Data is available online at University of Arizona ReData. Additional data is available upon reasonable request to the authors.

https://lib.arizona.edu/research/data/redata

## Notes

### Competing Interest Statement

The authors have declared no competing interest.

### Funding Statement

Dr. Valencia was supported by an NIH K12 award. Additional support was provided by the Doris Duke Charitable Trust FUTURRE Grant awarded to Dr. Valencia.

## References

[1] Giaquinto AN, Sung H, Miller KD, Kramer JL, Newman LA, Minihan A, et al. Breast Cancer Statistics, 2022. CA Cancer J Clin 2022;72:524–41. 10.3322/caac.21754.

[2] Jatoi I, Sung H, Jemal A. The Emergence of the Racial Disparity in U.S. Breast-Cancer Mortality. New England Journal of Medicine 2022;386:2349–52. 10.1056/nejmp2200244.

[3] Yedjou CG, Sims JN, Miele L, Noubissi F, Lowe L, Fonseca DD, et al. Health and racial disparity in breast cancer. Breast Cancer Metastasis and Drug Resistance: Challenges and Progress 2019:31–49.

[4] Lorona NC, Malone KE, Li CI. Racial/ethnic disparities in risk of breast cancer mortality by molecular subtype and stage at diagnosis. Breast Cancer Res Treat 2021;190:549–58. 10.1007/s10549-021-06311-7.

[5] Rock CL, Thomson C, Gansler T, Gapstur SM, McCullough ML, Patel A V, et al. American Cancer Society guideline for diet and physical activity for cancer prevention. CA Cancer J Clin 2020;70:245–71.

[6] Nicholson WK, Silverstein M, Wong JB, Barry MJ, Chelmow D, Coker TR, et al. Screening for Breast Cancer: US Preventive Services Task Force Recommendation Statement. JAMA 2024;331:1918–30. 10.1001/jama.2024.5534.

[7] Britt KL, Cuzick J, Phillips KA. Key steps for effective breast cancer prevention. Nat Rev Cancer 2020;20:417–36.

[8] Abdelhadi O, Williams M, Yan A. Structural racism as a leading cause of racial disparities in breast cancer quality of care outcomes: a systematic review. Front Oncol 2025;15. 10.3389/fonc.2025.1562672.

[9] Boone-Heinonen J, Diez Roux A V., Kiefe CI, Lewis CE, Guilkey DK, Gordon-Larsen P. Neighborhood socioeconomic status predictors of physical activity through young to middle adulthood: The CARDIA study. Soc Sci Med 2011;72:641–9. 10.1016/j.socscimed.2010.12.013.

[10] Kind AJH, Buckingham WR. Making Neighborhood-Disadvantage Metrics Accessible — The Neighborhood Atlas. New England Journal of Medicine 2018;378:2456–8. 10.1056/nejmp1802313.

[11] Plascak JJ, Beyer K, Xu X, Stroup AM, Jacob G, Llanos AAM. Association between Residence in Historically Redlined Districts Indicative of Structural Racism and Racial and Ethnic Disparities in Breast Cancer Outcomes. JAMA Netw Open 2022;5:E2220908. 10.1001/jamanetworkopen.2022.20908.

[12] Krieger N, Wright E, Chen JT, Waterman PD, Huntley ER, Arcaya M. Cancer Stage at Diagnosis, Historical Redlining, and Current Neighborhood Characteristics: Breast, Cervical, Lung, and Colorectal Cancers, Massachusetts, 2001-2015. Am J Epidemiol 2020;189:1065–75. 10.1093/aje/kwaa045.

[13] Wright E, Waterman PD, Testa C, Chen JT, Krieger N. Breast Cancer Incidence, Hormone Receptor Status, Historical Redlining, and Current Neighborhood Characteristics in Massachusetts, 2005-2015. JNCI Cancer Spectr 2022;6. 10.1093/jncics/pkac016.

[14] Gomby GA. Reassessing HOLC redlining maps to support claims of environmental injustice. Proc Natl Acad Sci U S A 2022;119. 10.1073/pnas.2200211119.

[15] Fishback P, Rose J, Snowden KA, Storrs T. New Evidence on Redlining by Federal Housing Programs in the 1930s. J Urban Econ 2024;141. 10.1016/j.jue.2022.103462.

[16] U.S. Shelley v. Kraemer. vol. 334. 1948.

[17] Otero LR. La Calle: spatial conflicts and urban renewal in a southwest city. Tucson, AZ: University of Arizona Press; 2010.

[18] Rothstein R. The Color of Law: A Forgotten History of How Our Government Segregated America. Norton; 2017.

[19] Melcher M. “This Is Not Right”: Rural Arizona Women Challenge Segregation and Ethnic Division, 1925-1950. Frontiers: A Journal of Women Studies 1999;20:190–214.

[20] López FA. Asset Pedagogies in Latino Youth Identity and Achievement: Nurturing Confianza. Taylor & Francis; 2017.

[21] Genesis L. TUSD board gains full control as desegregation case closes. Arizona Daily Star 2022.

[22] Jurjevich JR, Korgaonkar Y, Kollen C, Wilshin L, Holloway A. Mapping Racist Covenants. Mapping Racist Covenants 2023.

[23] U.S. Census Bureau. HISPANIC OR LATINO, AND NOT HISPANIC OR LATINO BY RACE. Decennial Census, DEC Demographic and Housing Characteristics, Table P9 2020.

[24] Pima County Health Department. Well Woman HealthCheck. Breast and Cervical Screening 2025.

[25] Arizona Department of Health Services. Arizona WIC and SNAP Locations 2024.

[26] Arizona Farmers Market Nutrition Program. Arizona Farmers Market Nutrition Program Find a Market 2025.

[27] Pima County. Parks and Recreational Areas 2020.

[28] Hammond E. Children’s Equity Nature Index (CENI): An Analysis of Young Children’s Equitable Access to Public Green Spaces in Urban Pima County, Arizon. 2024.

[29] Phillips A, Plastara D, Khan AZ, Canters F. Integrating public perceptions of proximity and quality in the modelling of urban green space access. Landsc Urban Plan 2023;240. 10.1016/j.landurbplan.2023.104875.

[30] Flanagan BE, Gregory EW, Hallisey EJ, Heitgerd JL, Lewis B. A Social Vulnerability Index for Disaster Management. J Homel Secur Emerg Manag 2020;8. 10.2202/1547-7355.1792.

[31] Centers for Disease Control and Prevention/ Agency for Toxic Substances and Disease Registry/ Geospatial Research A and SP. CDC/ATSDR Social Vulnerability Index 2022 2025.

[32] Bauer C, Zhang K, Xiao Q, Lu J, Hong YR, Suk R. County-Level Social Vulnerability and Breast, Cervical, and Colorectal Cancer Screening Rates in the US, 2018. JAMA Netw Open 2022;5. 10.1001/jamanetworkopen.2022.33429.

[33] Stuart CM, Dyas AR, Bronsert MR, Velopulos CG, Henderson WG, Schulick RD, et al. The association between social vulnerability and oncologic stage and treatment in the United States. Surgical Oncology Insight 2024;1. 10.1016/j.soi.2024.100044.

[34] Councell KA, Polcari AM, Nordgren R, Skolarus TA, Benjamin AJ, Shubeck SP. Social vulnerability is associated with advanced breast cancer presentation and all-cause mortality: a retrospective cohort study. Breast Cancer Research 2024;26. 10.1186/s13058-024-01930-6.

[35] Nguyen AT, Li RA, Ontiveros NC, Adam TH, Hansen N, Galiano RD. Utility of the Social Vulnerability Index in Addressing Breast Cancer Disparities: A Meta-Analysis. J Surg Oncol 2025. 10.1002/jso.70080.

[36] Flanagan BE, Gregory EW, Hallisey EJ, Heitgerd JL, Lewis B. A Social Vulnerability Index for Disaster Management. J Homel Secur Emerg Manag 2020;8. 10.2202/1547-7355.1792.

[37] Echeverría SE, Borrell LN, Brown D, Rhoads G. A local area analysis of racial, ethnic, and neighborhood disparities in breast cancer staging. Cancer Epidemiology Biomarkers and Prevention 2009;18:3024–9. 10.1158/1055-9965.EPI-09-0390.

[38] Goel N, Hernandez AE, Mazul A. Neighborhood Disadvantage and Breast Cancer-Specific Survival in the US. JAMA Netw Open 2024;7:E247336. 10.1001/jamanetworkopen.2024.7336.

[39] Moazzam Z, Woldesenbet S, Endo Y, Alaimo L, Lima HA, Cloyd J, et al. Association of Historical Redlining and Present-Day Social Vulnerability with Cancer Screening. J Am Coll Surg 2023;237:454–64. 10.1097/XCS.0000000000000779.

[40] Mah JC, Penwarden JL, Pott H, Theou O, Andrew MK. Social vulnerability indices: a scoping review. BMC Public Health 2023;23. 10.1186/s12889-023-16097-6.

[41] Councell KA, Polcari AM, Nordgren R, Skolarus TA, Benjamin AJ, Shubeck SP. Social vulnerability is associated with advanced breast cancer presentation and all-cause mortality: a retrospective cohort study. Breast Cancer Research 2024;26. 10.1186/s13058-024-01930-6.

[42] Bauer C, Zhang K, Xiao Q, Lu J, Hong YR, Suk R. County-Level Social Vulnerability and Breast, Cervical, and Colorectal Cancer Screening Rates in the US, 2018. JAMA Netw Open 2022;5. 10.1001/jamanetworkopen.2022.33429.

[43] Valencia CI, Curtis MG. Challenging the Urban-Rural Divide: Latino Parental Perceptions of the Built Environment 2025. 10.21203/rs.3.rs-6784739/v1.

[44] City of Tucson Parks and Recreation Department. City of Tucson Parks and Recreation System Master Plan. Tucson: 2016.

[45] Wiener CS, Carrel M, Zahnd WE. Iowa spatial accessibility to screening mammography is improving, but utilization among privately insured women remains stable, 2016–2022. Health Place 2025;94. 10.1016/j.healthplace.2025.103488.

[46] Arizona Healthcare Cost Containment System - Arizona Department of Health Services. AHCCCS Insights: Data to Inform Decision-Making. AHCCCS Insights: Data to Inform Decision-Making 2025.

[47] US Census Bureau. Population - Arizona. 1930.

[48] Hernandez AE, Borowsky PA, Nahodyl L, Pinheiro PS, Kobetz EN, Antoni MH, et al. A Neighborhood-Level Hispanic Paradox: The Interaction among Hispanic Density, Neighborhood Disadvantage, and Survival in Patients with Breast Cancer. Cancer Epidemiology Biomarkers and Prevention 2025;34:483–90. 10.1158/1055-9965.EPI-24-1242.

[49] U.S. Census Bureau - U.S. Department of Commerce. Commuting Characteristics by Sex. American Community Survey, ACS 5-Year Estimates Subject Tables, Table S0801 2025.

[50] Sprague BL, Ahern TP, Herschorn SD, Sowden M, Weaver DL, Wood ME. Identifying key barriers to effective breast cancer control in rural settings. Prev Med (Baltim) 2021;152. 10.1016/j.ypmed.2021.106741.

[51] Webster JL, Goldstein ND, Rowland JP, Tuite CM, Siegel SD. A catchment and location-allocation analysis of mammography access in Delaware, US: implications for disparities in geographic access to breast cancer screening. Breast Cancer Research 2023;25. 10.1186/s13058-023-01738-w.

[52] Molina Y, Lee E, Piñeros J, Ries D, Gonzalez J, Lofton S, et al. Characterizing the catchment area and identifying scientific priorities with communities: An example from the University of Illinois Cancer Center. Preventive Oncology & Epidemiology 2024;2. 10.1080/28322134.2024.2381819.

[53] Geraghty EM. Enhancing cancer care, research, and engagement through geographic context: The role of geographic information systems in defining and serving cancer center catchment areas. Preventive Oncology & Epidemiology 2024;2. 10.1080/28322134.2024.2379437.

[54] Hashtarkhani S, Zhou Y, Kumsa FA, White-Means S, Schwartz DL, Shaban-Nejad A. Analyzing Geospatial and Socioeconomic Disparities in Breast Cancer Screening Among Populations in the United States: Machine Learning Approach. JMIR Cancer 2025;11. 10.2196/59882.

